# Early Anti-SARS-CoV-2 Convalescent Plasma in Patients Admitted for COVID-19: A Randomized Phase II Clinical Trial

**DOI:** 10.1101/2020.09.17.20196212

**Authors:** María Elvira Balcells, Luis Rojas, Nicole Le Corre, Constanza Martínez-Valdebenito, María Elena Ceballos, Marcela Ferrés, Mayling Chang, Cecilia Vizcaya, Sebastián Mondaca, Álvaro Huete, Ricardo Castro, Mauricio Sarmiento, Luis Villarroel, Alejandra Pizarro, Patricio Ross, Jaime Santander, Barbara Lara, Marcela Ferrada, Sergio Vargas-Salas, Carolina Beltrán-Pavez, Ricardo Soto-Rifo, Fernando Valiente-Echeverría, Christian Caglevic, Mauricio Mahave, Carolina Selman, Raimundo Gazitúa, José Luis Briones, Franz Villarroel-Espindola, Carlos Balmaceda, Manuel A. Espinoza, Jaime Pereira, Bruno Nervi

**Affiliations:** Department of Infectious Diseases. School of Medicine. Pontificia Universidad Católica de Chile; Department of Internal Medicine. School of Medicine. Pontificia Universidad Católica de Chile; Program of Pharmacology and Toxicology. School of Medicine. Pontificia Universidad Católica de Chile; Department of Pediatric Infectious Diseases and Immunology. School of Medicine. Pontificia Universidad Católica de Chile; Diagnostic Virology Laboratory, Red de Salud UC CHRISTUS; Department of Hematology and Oncology. School of Medicine. Pontificia Universidad Católica de Chile; Department of Radiology. School of Medicine. Pontificia Universidad Católica de Chile; Department of Intensive Care Medicine. School of Medicine. Pontificia Universidad Católica de Chile; Department of Public Health. School of Medicine. Pontificia Universidad Católica de Chile; Department of Psychiatry. School of Medicine. Pontificia Universidad Católica de Chile; Emergency Medicine Section. School of Medicine. Pontificia Universidad Católica de Chile; Clinical Research Center. School of Medicine. Pontificia Universidad Católica de Chile; Laboratory of Molecular and Cellular Virology, Virology Program, Institute of Biomedical Sciences, Faculty of Medicine, Universidad de Chile. HIV/AIDS Work Group, Faculty of Medicine, Universidad de Chile; Instituto Oncológico Fundación Arturo López Pérez (FALP) Santiago, Chile; Translational Medicine Research Laboratory. Fundación Arturo López Pérez (FALP) Santiago, Chile; Health Technology Assessment Unit, Clinical Research Center, School of Medicine, Pontificia Universidad Católica de Chile

**Author notes:** Corresponding author: María Elvira Balcells. Department of Infectious Diseases. School of Medicine. Pontificia Universidad Católica de Chile. Address: Diagonal Paraguay 362, Of.617. Santiago, Chile. Co-first authorship (María Elvira Balcells, Luis Rojas).

**Keywords:** plasma, convalescent plasma, COVID-19, SARS-CoV-2

## Abstract

**Background:** Convalescent plasma (CP), despite limited evidence on its efficacy, is being widely used as a compassionate therapy for hospitalized patients with COVID-19. We aimed to evaluate the efficacy and safety of early CP therapy in COVID-19 progression.

**Methods:** Open-label, single-center, randomized clinical trial performed in an academic center in Santiago, Chile from May 10, 2020, to July 18, 2020, with final follow-up August 17, 2020. The trial included patients hospitalized within the first 7 days of COVID-19 symptoms onset, presenting risk factors for illness progression and not on mechanical ventilation. The intervention consisted in immediate CP (early plasma group) versus no CP unless developing pre-specified criteria of deterioration (deferred plasma group). Additional standard treatment was allowed in both arms. The primary outcome was a composite of mechanical ventilation, hospitalization for >14 days or death. Key secondary outcomes included: time to respiratory failure, days of mechanical ventilation, hospital length-of-stay, mortality at 30 days, and SARS-CoV-2 RT-PCR clearance rate.

**Results:** Of 58 randomized patients (mean age, 65.8 years, 50% male), 57 (98.3%) completed the trial. A total of 13 (43.3%) participants from the deferred group received plasma based on clinical aggravation. We found no benefit in the primary outcome (32.1% vs 33.3%, OR 0.95, 95% CI 0.32-2.84, p>0.99) in the early versus deferred CP group. In-hospital mortality rate was 17.9% vs 6.7% (OR 3.04, 95% CI 0.54-17.2, p=0.25), mechanical ventilation 17.9% vs 6.7% (OR 3.04, 95% CI 0.54-17.2, p=0.25), and prolonged hospitalization 21.4% vs 30% (OR 0.64, 95%CI, 0.19-2.1, p=0.55) in early versus deferred CP group, respectively. Viral clearance rate on day 3 (26% vs 8%, p=0.20) and day 7 (38% vs 19%, p=0.37) did not differ between groups. Two patients experienced serious adverse events within 6 or less hours after plasma transfusion.

**Conclusion:** Immediate addition of CP therapy in early stages of COVID-19 -compared to its use only in case of patient deterioration-did not confer benefits in mortality, length of hospitalization or mechanical ventilation requirement.

**Clinical Trials Registration:** NCT04375098

## Introduction

The SARS-CoV-2 pandemic has left over 24 million contagions and 833,000 deaths by August 29, 2020[1]. Different case-series have shown an intensive care unit (ICU) admission rate between 5 and 16%, and a case fatality rate near 1-4%, with a direct relationship with age and comorbidities[2–4]. To date, there is no validated treatment nor immunization against SARS-CoV-2. A promising alternative is immune plasma from convalescent patients[5]. This strategy has been used with some success in other viral diseases with important lethality such as hantavirus, influenza, SARS-CoV, and MERS-CoV infections[6–9].

The use of convalescent plasma for COVID-19 was reported early in this pandemic. The initial case-series studies suggested faster clinical recovery, viral clearance and radiologic improvement, although the lack of a controlled group limited the accurate interpretation of these results[10–12]. Subsequently, a preliminary report of a matched controlled study showed convalescent plasma improved survival for non-intubated patients[13]. However, the first two randomized controlled trials showed no clear clinical benefit, and furthermore, one of these trials was stopped early due to concerns when finding high pre-existing SARS-CoV-2 neutralizing antibody titers in receptors before transfusion[14,15]

Considering that COVID-19 likely involves at least two phases — an early phase in which viral replication is a component of tissue injury and a later phase in which a dysregulated and pro-inflammatory immune response leads the damage — the most useful therapeutic window for convalescent plasma administration is currently unknown[16]. Indeed, the lack of efficacy of previous studies has been attributed to a late timing of plasma administration in the disease’s course. This hypothesis is consistent with the recent finding of lower mortality for patients receiving convalescent plasma within the first 3 days after COVID-19 diagnosis in a large uncontrolled study[17].

The objective of this study was to assess the efficacy and safety of convalescent plasma therapy in reducing disease progression, complications, and death in patients in the early phase of COVID-19.

## Patients and Methods

This study consisted of a randomized, controlled, open-label phase II trial done in a single Chilean medical center in Santiago, Chile. Patients were randomized from May 10, 2020, to July 18, 2020, with final follow-up August 17, 2020.

### Inclusion criteria

Inclusion criteria were the following: (1) patients over 18 years old; (2) hospitalized, COVID-19 symptoms present at enrollment and confirmed with a positive SARS-CoV-2 real-time polymerase chain reaction (RT-PCR) in nasopharyngeal swab or, pending PCR result but imaging consistent with COVID-19 pneumonia and confirmed COVID-19 close-contact; (3) ≤ 7 days from COVID-19 symptoms onset to enrollment; (4) a CALL score ≥ 9 points at enrollment (predicts high-risk of progression into respiratory failure, based on age, comorbidities, lactate dehydrogenase (LDH) and lymphocyte count)[18]; (5) Eastern Cooperative Oncology Group (ECOG) performance status before SARS-CoV-2 infection 0-2.

### Exclusion criteria

Exclusion criteria were the following: (1) PaO_2_/FiO_2_<200 or need for mechanical ventilation at enrollment; (2) coinfection with other relevant respiratory pathogens on admission; (3) pregnancy or lactation; (4) known IgA Nephropathy or IgA deficiency; (5) previous immunoglobulin or plasma administration within the last 60 days; (6) previous severe transfusion reactions; (7) do not resuscitate indication; (8) participating in another COVID-19 interventional study; and (9) patients that under investigator criteria had any condition that made them unsuitable for study participation.

### Convalescent plasma donation protocol

Plasma was obtained from volunteer subjects who had recovered from COVID-19, having been asymptomatic for at least 28 days, with a negative SARS-CoV-2 RT-PCR both in nasopharyngeal swab and in plasma, and anti-SARS-CoV-2 (S1) IgG titers ≥ 1:400 (ELISA Euroimmun®). Donor plasma was tested for standard infectious diseases before administration and extracted plasma was immediately frozen at -20°C according to standard national safety measures[19].

### Randomization and intervention

Eligible patients were randomly assigned via computer-generated numbering by a block randomization sequence into two groups: early or deferred plasma transfusion. Randomization was done by an independent member, and the sequence was concealed to study investigators.

The early plasma group received the first plasma unit at enrollment. The deferred plasma group received convalescent plasma only if a pre-specified worsening respiratory function criterion was met during hospitalization (Pa02/Fi02 <200) or if the patient still required hospitalization for symptomatic COVID-19 >7 days after enrollment (Figure 1).

**Figure 1.**
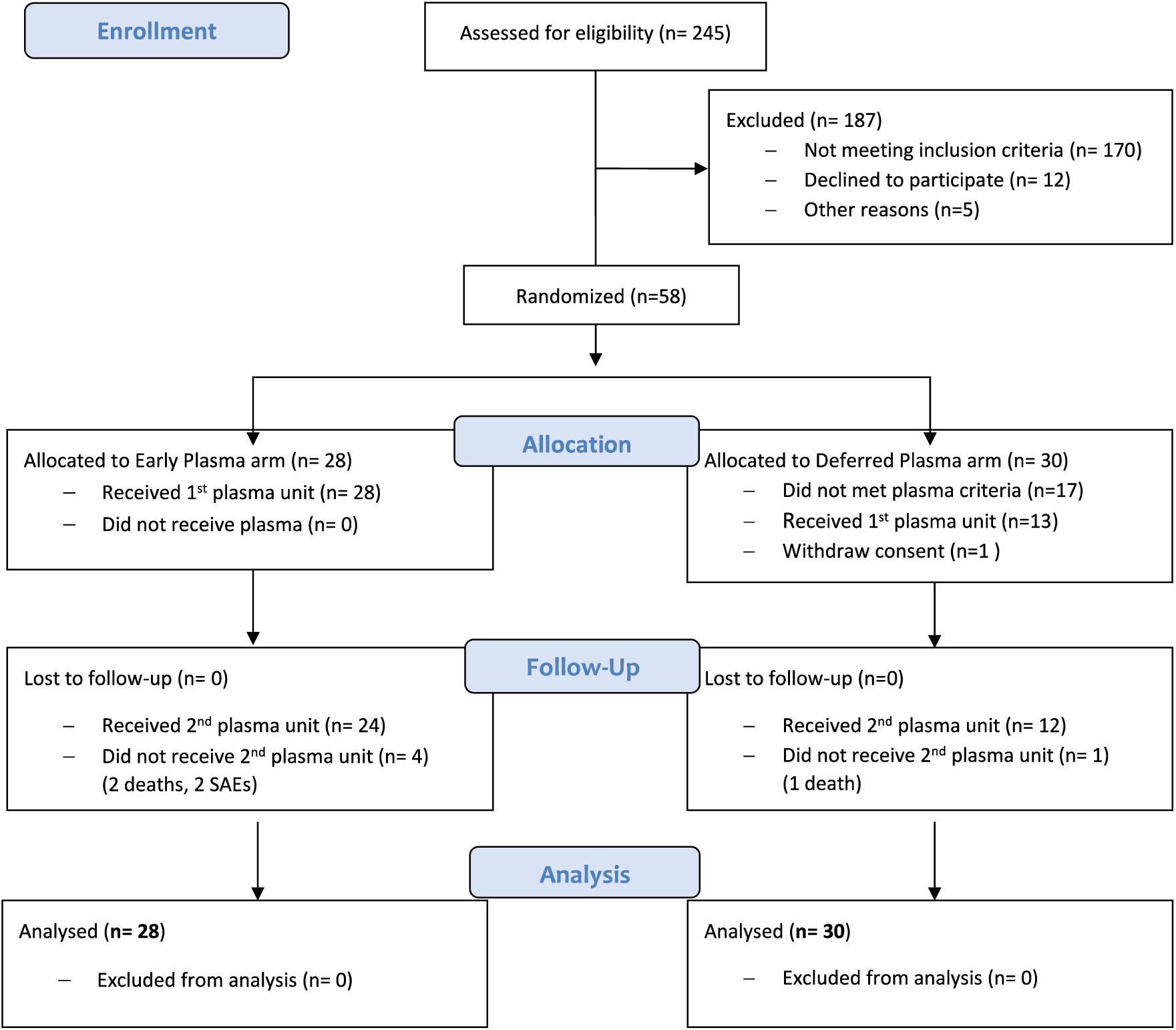
Study Flow Diagram. Patient enrollment and treatment assignment

Transfusions considered a total of 400 mL of ABO compatible convalescent plasma, infused as two 200 mL units, each separated by 24 hours. In both groups, cointerventions, including antibiotics, antivirals, heparin thromboprophylaxis, and immunomodulators, were allowed based on the hospital protocols.

### Outcomes

The primary outcome was a composite of mechanical ventilation, hospitalization >14 days or in-hospital death.

Secondary outcomes included: days of mechanical ventilation, days of high-flow nasal cannula (HFNC), days of oxygen requirement, time to respiratory failure development (PaO_2_/FiO_2_<200), the severity of multiple organ dysfunction (by SOFA score) at day 3 and 7; days in ICU or intermediate care unit, hospital length of stay, and mortality at 30 days. The kinetics of inflammatory biomarkers, including total lymphocyte count, C-reactive protein (CRP), procalcitonin, LDH, D-dimer, ferritin and IL-6 were determined on days 0, 3 and 7; and SARS-CoV-2 RT-PCR in nasopharyngeal swab on days 3 and 7.

Radiological outcomes included the comparison of infiltrates progression on chest CT scans on enrollment and day 5, based on COVID-19 pneumonia severity scores[20–23]. For the combined analysis with portable chest X-rays, a blinded thoracic radiologist expert categorized images as “progression” vs “stable or improved”.

Also, pre-planned analyses of baseline neutralizing antibodies (NAbs) titers, and anti-SARS-CoV-2 IgG titers were determined in participants from the early plasma group and in the subset of participants from the deferred plasma group who had not yet received plasma on days 0, 3 and 7.

Analysis of the primary outcome and clinical secondary outcomes was performed by intention-to-treat (ITT). Laboratory and radiology secondary outcomes were analyzed by modified-ITT, excluding a patient who withdrew consent before any intervention.

Safety outcomes were evaluated in all participants.

### Anti-SARS-CoV-2 IgG ELISA

For specific IgG enzyme-linked immunosorbent assays (ELISA) we used the commercial kit CE-marked Euroimmun (Lübeck, Germany, # EI 2606-9601 G), which uses the S1-domain of spike protein of SARS-CoV-2 as antigen. Fresh or thawed serum samples were first diluted at 1:101, then 2-fold serial dilutions were done until 1:6400. Immunoreactivity was measured at an optical density (OD) of 450 nm. Results were expressed according to the manufacturer. The end-point dilution for each sample was determined as the final dilution where the OD ratio (patient/calibrator) was ≥1.1. Seroconversion was defined as seronegative at baseline and seropositive after 3 or 7 days or, a 4-fold increase in end-point dilution titer from the baseline.

### Neutralizing antibody titers assay

Anti-SARS-CoV-2 NAbs were measured in serum samples using an HIV-1 backbone expressing firefly luciferase as a reporter gene and pseudotyped with the SARS-CoV-2 Spike glycoprotein[24,25]. Given the results obtained in an ongoing study with pre-pandemic sera in Chilean population, 1:160 was defined as the screening dilution (Beltrán-Pavez, *et al*. Manuscript in preparation). Samples with a neutralizing activity of at least 50% at a 1:160 dilution were considered positive and used to perform titration curves and 50% neutralization titer (ID50) calculations. Determination of the ID50 was performed using a 4-parameter nonlinear regression curve fit measured as the percent of neutralization determined by the difference in average relative light units (RLU) between test wells and pseudotyped virus controls. In order to perform the ID50 calculations, the lack of fit test had a p-value > 0.1. The top values were constrained to 100 and the bottom values were set to 0 (Beltrán-Pavez C, *et al*. Manuscript in preparation).

### Statistical Analysis

Sample size was calculated with a power of 80% and a statistical significance of 5% for an absolute risk reduction of 35% in the primary outcome based on a previous report of convalescent plasma administration in the early stage of AH1N1 influenza[26]. The final calculated sample size was of 29 individuals per group (total n=58).

The primary binary outcomes were assessed through chi-squared tests or Fisher’s exact test when appropriate, and odds ratios (ORs) are presented together with 95% CI and p-values. Results of the main analysis are presented as crude analysis. In addition, we adjusted for age and SOFA score at enrolment, as fixed (individual-level) effects, using logistic regression. For secondary outcomes, binary variables were analyzed using logistic regression. Numerical variables were examined using generalized linear models with log link function and gamma family function. For those variables with high number of zeros, we used a zero-inflated negative binomial model because it showed better goodness of fit compared with other zero-inflated models according to the Akaike information criterion. Treatment effects estimates, crude and adjusted by age and SOFA, are presented as exponentiated coefficients, i.e., ORs and incidence rate ratios (IRRs), respectively, with their corresponding 95%CI. In those cases where asymptotic assumptions did not hold, crude estimates were analyzed with Fisher’s exact test for categorical variables and Wilcoxon-rank-sum test for continuous variables. To test differences between Kaplan-Meier estimates in survival analysis, we used the Log-Rank test.

For paired CT scans scores analysis, we used Wilcoxon matched-pairs signed-rank test.

Statistical significance was defined using a 2-sided significance level of α = .05. The statistical analysis was performed by an investigator who was blind to the study group allocation. Analyses were done with R version 3.6.3 and figures with GraphPad Prism version 8.4.3 software.

### Ethics

This study was approved by the Institutional Review Board of the Pontificia Universidad Católica de Chile. Written informed consent was solicited from each patient or their legal representatives.

## Results

### Study population

Of the 245 patients diagnosed with COVID-19 and evaluated for eligibility, a total of 58 patients were enrolled and included in the ITT analysis. The mean age was 65.8 years (range: 27-92), and 50% were women. The median interval between symptom onset to randomization was 6 days (IQR 4-7). All patients had SARS-CoV-2 infection confirmed by RT-PCR in nasopharyngeal swab. Baseline characteristics of participants are described in Table 1.

**Table 1.** Baseline demographics and clinical characteristics of study participants

|  | Early plasma group<br>(n= 28) | Deferred plasma group<br>(n=30) | P-value |
| --- | --- | --- | --- |
| Age (years), mean (range) | 64.3 (33-92) | 67.1 (27-91) | 0.42 |
| Male sex, N° (%) | 15 (53.6) | 14 (46.7) | 0.79 |
| Blood group A, N°/total (%) | 7/28 (25.0) | 7/27 (25.9) | >0.99 |
| Blood group O, N°/total (%) | 19/28 (67.9) | 14/27 (51.8) | 0.28 |
| Obesity (BMI >30), N° (%) | 3 (10.7) | 4 (13.3) | >0.99 |
| Diabetes mellitus, N° (%) | 10 (35.7) | 11(36.7) | >0.99 |
| Hypertension, N° (%) | 17 (60.7) | 22 (73.3) | 0.40 |
| Cerebrovascular disease, N° (%) | 3 (10.7) | 0 (0) | 0.11 |
| Cancer, N° (%) | 1 (3.6) | 3 (10.0) | 0.61 |
| Immunosuppressants, N° (%) | 4 (14.3) | 3 (10.0) | 0.70 |
| Chronic renal failure N° (%) | 2 (7.1) | 3 (10.0) | >0.99 |
| Chronic liver disease , N° (%) | 3 (10.7) | 0 (0.0) | 0.11 |
| Asthma, N° (%) | 1 (3.6) | 2 (6.7) | >0.99 |
| Days since COVID symptoms initiation, median (IQR) | 5 (4-7) | 6 (4-7) | 0.70 |
| CALL score at enrollment<sup>a</sup>, median (IQR) | 10.5 (10-12) | 10.0 (10-12) | 0.81 |
| SOFA score at enrollment<sup>b</sup>, median (IQR) | 2.0 (2.0-4.0) | 2.0 (2.0-2.0) | 0.21 |
| O<sub>2</sub> requirement at enrollment, N° (%) | 23 (82.1) | 23 (76.7) | 0.75 |
| FiO<sub>2</sub> requirement at enrollment, median (IQR) | 0.28 (0.22-0.30) | 0.24 (0.21-0.28) | 0.30 |
| PaO<sub>2</sub>/FiO<sub>2</sub> at enrollment, median (IQR) | 260.7 (211-316) | 260.7 (222-308) | 0.46 |
| Lung infiltrates in CT scan or Chest X-ray, N° (%) | 28 (100.0) | 29 (96.7) | >0.99 |
| **Baseline chest CT severity score:** |  |  |  |
| - Criteria 1<sup>c</sup><br>Median score (IQR) (N°) | 18.0 (11.5-26.0)<br>(25) | 14.0 (10.0-19.0)<br>(26) | 0.27 |
| - Criteria 2<sup>d</sup><br>Median score (IQR) (N°) | 20.0 (15.5-28.0)<br>(25) | 18.0 (15.8-22.5)<br>(26) | 0.19 |
| - Criteria 3<sup>e</sup><br>Median score (IQR) (N<sup>o</sup>) | 13.0 (10.0-17.5)<br>(25) | 11.0 (9.0-13.0)<br>(26) | 0.09 |
| - Severe pneumonia on CT (severity score >19, criteria 2<sup>d</sup>) N<sup>o</sup>/total (%) | 14/25 (56.0) | 09/26 (34.6) | 0.16 |
| - Severe pneumonia on CT (severity score ≥ 12, criteria 3<sup>e</sup>) N<sup>o</sup>/total (%) | 17/25 (68.0) | 12/26 (46.1) | 0.16 |
| **Other pharmacological interventions for COVID-19 during hospitalization:** |  |  |  |
| - Steroids,<br>N<sup>o</sup> (%) | 16 (57.1) | 14 (46.7) | 0.44 |
| - IL6 blocker (tocilizumab) ,<br>N<sup>o</sup> (%) | 1 (3.6) | 1 (3.3) | >0.99 |
| - Hydroxicloroquine ,<br>N<sup>o</sup> (%) | 2 (7.14) | 5 (16.7) | 0.42 |
| - Lopinavir/ritonavir,<br>N<sup>o</sup> (%) | 1 (3.6) | 0 (0.0) | 0.48 |
| - Thromboprophylaxis<sup>f</sup>,<br>N<sup>o</sup> (%) | 25 (89.3) | 23 (76.7) | 0.30 |
| - Anticoagulation<sup>f</sup>,<br>N<sup>o</sup> (%) | 2 (7.14) | 6 (20.0) | 0.25 |
| a) CALL score (risk of progression into respiratory failure, based on age, comorbidities, LDH and lymphocyte count). Ref. Ji D et al.[18]<br>b) Sequential Organ Failure Assessment (SOFA) Score<br>c) Chest CT severity Score 1. Ref. Zhou et al[20]<br>d) Chest CT severity Score 2. Ref. Yang et al[23]<br>e) Chest CT severity Score 3. Ref. Pan et al[21,22,36]<br>f) Unfractionated or low-molecular weight heparin |  |  |  |

All participants (n=28) from the early plasma group received a 1^st^ plasma unit on the day of enrollment and 24 (86%) received a 2^nd^ unit 24 hours later. Reasons for not receiving the 2^nd^ unit were death (n=2) or a serious adverse event (SAE) after the first plasma unit administration (n=2).

A total of 13 participants (43.3%) from the deferred plasma group received plasma at a median time of 3 days from enrollment (IQR 1-5), based on respiratory failure development (n=12) or persistent symptomatic COVID-19 beyond 7 days after enrollment (n=1).

### Primary Outcome

There was no significant difference between the early and deferred plasma group in the composite primary outcome: 32.1% (9/28) in the early plasma group vs 33.3% (10/30) in the deferred plasma group (OR 0.95, 95%CI 0.32-2.84). When the outcome was disaggregated, the differences were 17.9% (5/28) vs 6.7% (2/30) (OR 3.04, 95%CI 0.54-17.2), for in-hospital death, 17.9% (5/28) vs 6.7% (2/30) (OR 3.04, 95% CI 0.54-17.2), for mechanical ventilation, and 21.4% (6/28) vs 30% (9/30) (OR 0.64, 95%CI 0.19-2.1) for hospitalization >14 days, in the early versus deferred plasma group, respectively (Table 2).

**Table 2.** Primary and main secondary clinical outcomes

|  | Early plasma group<br>(n=28) | Deferred plasma group<br>(n=30) | P-value<sup>a</sup> | Crude Effect estimate<br>(95%CI) | Adjusted - Effect estimate (95%CI) |
| --- | --- | --- | --- | --- | --- |
| **Primary clinical outcomes** |  |  |  |  |  |
| - Composite outcome (death, mechanical ventilation and/or hospital stay >14 days), N<sup>o</sup>/total (%) | 9/28 (32.1) | 10/30 (33.3) | >0.99 | OR 0.95 (0.32-2.84) | OR 0.67 (0.14-3.31) |
| - Mechanical ventilation, N<sup>o</sup>/total (%) | 5/28 (17.9) | 2/30 (6.7) | 0.25 | OR 3.04 (0.54-17.2) | OR 2.98 (0.41-21.57) |
| - Death, N<sup>o</sup>/total (%) | 5/28 (17.9) | 2/30 (6.7) | 0.25 | OR 3.04 (0.54-17.2) | OR 4.22 (0.33-53.57) |
| - Hospitalization >14 days, N<sup>o</sup>/total (%) | 6/28 (21.4) | 9/30 (30.0) | 0.55 | OR 0.64 (0.19-2.1) | OR 0.51 (0.13-2.05) |
| **Secondary clinical outcomes:** |  |  |  |  |  |
| - 30-day mortality, N<sup>o</sup>/total (%) | 5/28 (17.9) | 2/30 (6.7) | 0.25 | OR 3.04 (0.54-17.2) | OR 4.22 (0.33-53.57) |
| - Progression into respiratory failure<sup>b</sup>, N<sup>o</sup>/total (%) | 13/28 (46.4) | 12/30 (40.0) | 0.79 | OR 1.30 (0.46-3.68) | OR 1.46 (0.43 -4.66) |
| - Total days of mechanical ventilation requirement, median (IQR) | 0.0 (0.0-0.0) | 0.0 (0.0-0.0) | 0.23 | IRR 1.68 (0.30 -9.42) | IRR 4.78 (2.20 -10.40) |
| - Total days of HFNC<sup>d</sup> requirement, median (IQR) | 0.0 (0.0-2.5) | 0.0 (0.0-2.0) | 0.75 | IRR 0.70 (0.35-1.43) | IRR 0.65 (0.35 - 1.30) |
| - Total days of oxygen requirement, median (IQR) | 6.0 (3.0-12.0) | 7.0 (2.0-16.0) | 0.95 | IRR 0.90 (0.53-1.53) | IRR 1.07 (0.64 -1.78) |
| -Total days of intensive and/or intermediate care requirement, median (IQR) | 2.5 (0.0-8.25) | 0.0 (0.0-8.5) | 0.44 | IRR 0.69 (0.37-1.31) | IRR 0.68 (0.36-1.26) |
| - Total days of hospital stay, median (IQR) | 9.0 (5.0-12.0) | 8.0 (5.5-23.0) | 0.81 | IRR 0.78 (0.50-1.22) | IRR 0.86 (0.57-1.29) |
| - SOFA score<sup>c</sup> Day 3, median (IQR) | 2.0 (1.0-4.0) | 2.0 (1.0-3.0) | 0.73 | IRR 1.18 (0.78 -1.79) | IRR 1.12 (0.84 -1.48) |
| - SOFA score<sup>c</sup> Day 7, median (IQR) | 2.0 (2.0-4.0) | 3.0 (1.0-4.0) | 0.56 | IRR 1.29 (0.74-2.22) | IRR 0.98 (0.65 - 1.48) |
| a) p-value was calculated by Wilcoxon-rank-sum test or Fisher exact test.<br>b) Respiratory failure defined as PaO<sub>2</sub>/FiO<sub>2</sub><200<br>c) Sequential Organ Failure Assessment (SOFA) Score<br>d) HFNC: high-flow nasal cannula |  |  |  |  |  |

### Secondary Outcomes

A total of 46.4% of early plasma group participants progressed to severe respiratory failure (PaO2/FiO2<200) compared to 40% of patients from the deferred plasma group (OR 1.30, 95%CI 0.48-3.56) at a median time of 2.0 and 2.5 days from enrollment, respectively. No significant differences were noted in any of the other main secondary outcomes (Table 2). In the adjusted models, the total number of days in mechanical ventilation resulted higher in the early plasma than in the deferred plasma group (IRR 4.78, 95%CI 2.20 -10.40). Time to death and time to develop severe respiratory failure did not differ between both study groups (Figure 2).

**Figure 2.**
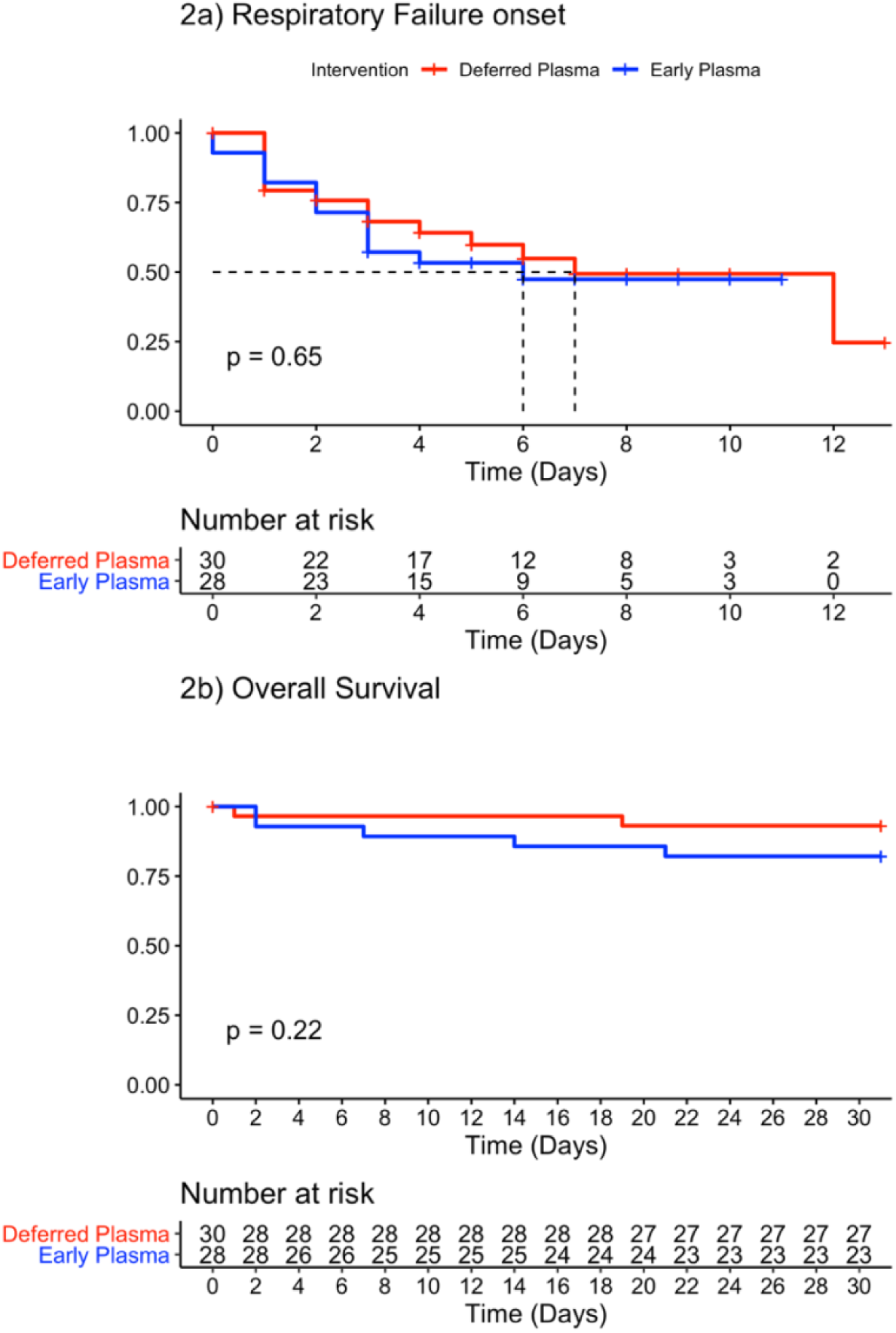
Time from enrollment to key secondary outcomes. A. Time from enrollment to severe respiratory failure development (PaO2/FiO2<200) in early plasma and deferred plasma groups. B. Time from enrollment to death in early plasma and deferred plasma groups.

No significant differences were found for CRP, IL-6, ferritin, LDH, D-dimer, procalcitonin and lymphocyte count levels on day 3 and 7 between study groups (Table 3). Similarly, the rate of SARS-CoV-2 negative PCR in nasopharyngeal swabs did not differ between early and deferred plasma groups on day 3 (26% vs 8%, p=0.20) nor on day 7 (38% vs 19%, p=0.37) (Figure 3).

**Table 3.** Laboratory outcomes

|  | Early plasma group<br>(n=28) | Deferred plasma group<br>(n=29) | P-value |
| --- | --- | --- | --- |
| **CRP<sup>a</sup> (mg/dl), median (IQR) (N<sup>o</sup>)** |  |  |  |
| - day 0 | 9.2 (5.1-15.2) (28) | 9.5 (4.1-16.1) (27) | 0.81 |
| - day 3 | 5,3 (2.1-12.8) (24) | 7.7 (3.15-12.5) (25) | 0.39 |
| - day 7 | 3.9 (1.2-5.6) (16) | 3.2 (1.2-9.2) (20) | 0.94 |
| **Lymphocyte count (x10<sup>9</sup>/L), median (IQR) (N<sup>o</sup>)** |  |  |  |
| - day 0 | 680 (490-910) (25) | 800 (530-1120) (27) | 0.46 |
| - day 3 | 760 (540-1190) (23) | 620 (520-840) (24) | 0.15 |
| - day 7 | 760 (450-1040) (17) | 910 (500-1260) (19) | 0.66 |
| **IL-6 (pg/ml), median (IQR) (N<sup>o</sup>)** |  |  |  |
| - day 0 | 46.3 (27.9-120.6) (28) | 53.3 (18.5-127.8) (25) | 0.88 |
| - day 3 | 23.6 (10.2-48.5) (24) | 20.8 (8-81.4) (26) | 0.86 |
| - day 7 | 42.9 (4.2-110.7) (17) | 14.8 (6-93.8) (19) | 1.00 |
| **Procalcitonin (ng/ml), median (IQR) (N<sup>o</sup>)** |  |  |  |
| - day 0 | 0.18 (0.09-0.73) (28) | 0.09 (0.08-0.20) (27) | 0.18 |
| - day 3 | 0.12 (0.09-0.31) (24) | 0.11 (0.08-0.24) (26) | 0.82 |
| - day 7 | 0.09 (0.08-0.21) (15) | 0.10 (0.08-0.14) (19) | 0.96 |
| **Ferritin (ng/ml), median (IQR) (N<sup>o</sup>)** |  |  |  |
| - day 0 | 881 (571-1714) (28) | 849 (599-1228) (28) | 0.58 |
| - day 3 | 1215 (589-1934) (24) | 1001 (572-2057) (26) | 0.78 |
| - day 7 | 1021 (520-1679) (16) | 974 (587-1556) (20) | 0.92 |
| **D-dimer (ng/ml), median (IQR) (N<sup>o</sup>)** |  |  |  |
| - day 0 | 1128 (658-1689) (28) | 926 (622-1133) (29) | 0.30 |
| - day 3 | 974 (614-1255) (24) | 835 (615-1326) (26) | 0.87 |
| - day 7 | 963 (723-2510) (15) | 948 (664-1987) (20) | 0.68 |
| **LDH (U/L), median (IQR) (N<sup>o</sup>)** |  |  |  |
| - day 0 | 317 (256-396) (28) | 299 (251-350) (27) | 0.35 |
| - day 3 | 331 (260-424) (24) | 320 (260-404) (25) | 0.78 |
| - day 7 | 290 (254-357) (17) | 317 (233-442) (20) | 0.58 |
| a) C- Reactive Protein |  |  |  |

**Figure 3.**
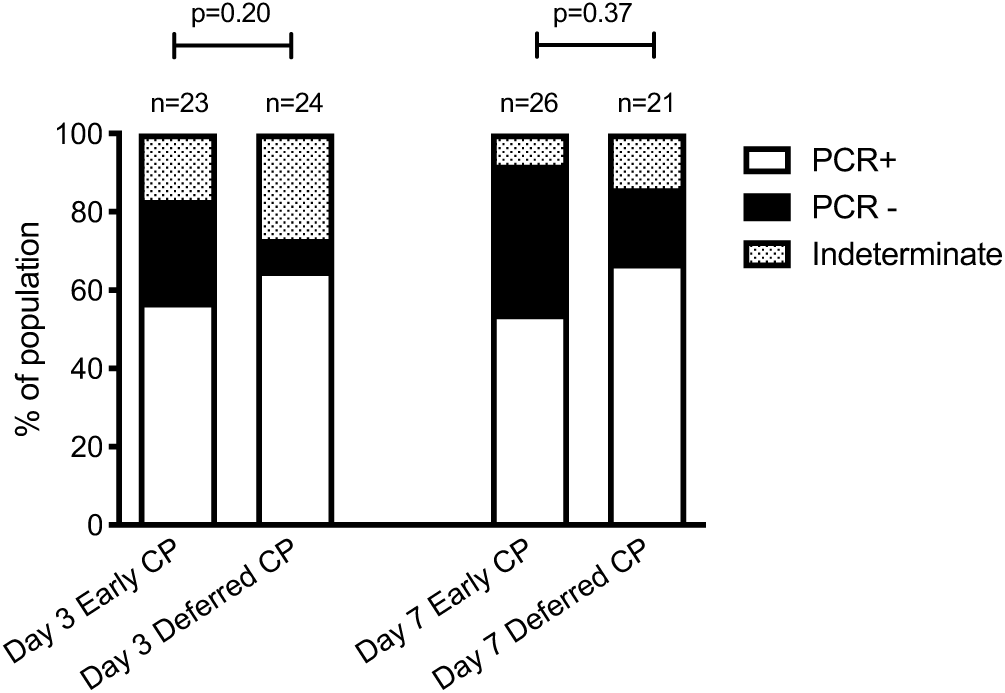
SARS-CoV-2 RT-PCR in nasopharyngeal samples. Each column represents the RT-PCR results obtained in patients from early plasma group and deferred plasma group. Above columns, the number of samples are indicated. The proportion of positive RT-PCR are represented in white, negative RT-PCR in black and an indeterminate result (Cycle Threshold ≥ 35) dashed.

The progression in the COVID pneumonia CT severity scores from baseline to day 5 was higher in the deferred than in the early plasma group (Figure 4). However, when the analysis also included subjects who had a chest X-ray instead of CT on the same scheduled days, the proportion of patients with progression in lung infiltrates did not differ between groups (OR 1.3, 95%CI 0.41-3.89) (Table 4).

**Table 4.** Radiological changes (from day 0 to day 5) for early and deferred plasma groups, based on expert radiologist criteria

| Blinded expert criteria<br>(combined CT and chest X-ray analysis) | Early plasma group<br>(n=21) | Deferred plasma group<br>(n=24) | Effect estimate<br>(95% CI) | P value |
| --- | --- | --- | --- | --- |
| - Stable or improved<br>Nº(%) | 10 (47.6) | 13 (54.2) | OR 1.30<br>(95%CI 0.41- 3.89) | 0.77 |
| - Progressed (worse)<br>Nº(%) | 11 (52.4) | 11 (45.8) |  |  |

**Figure 4.**
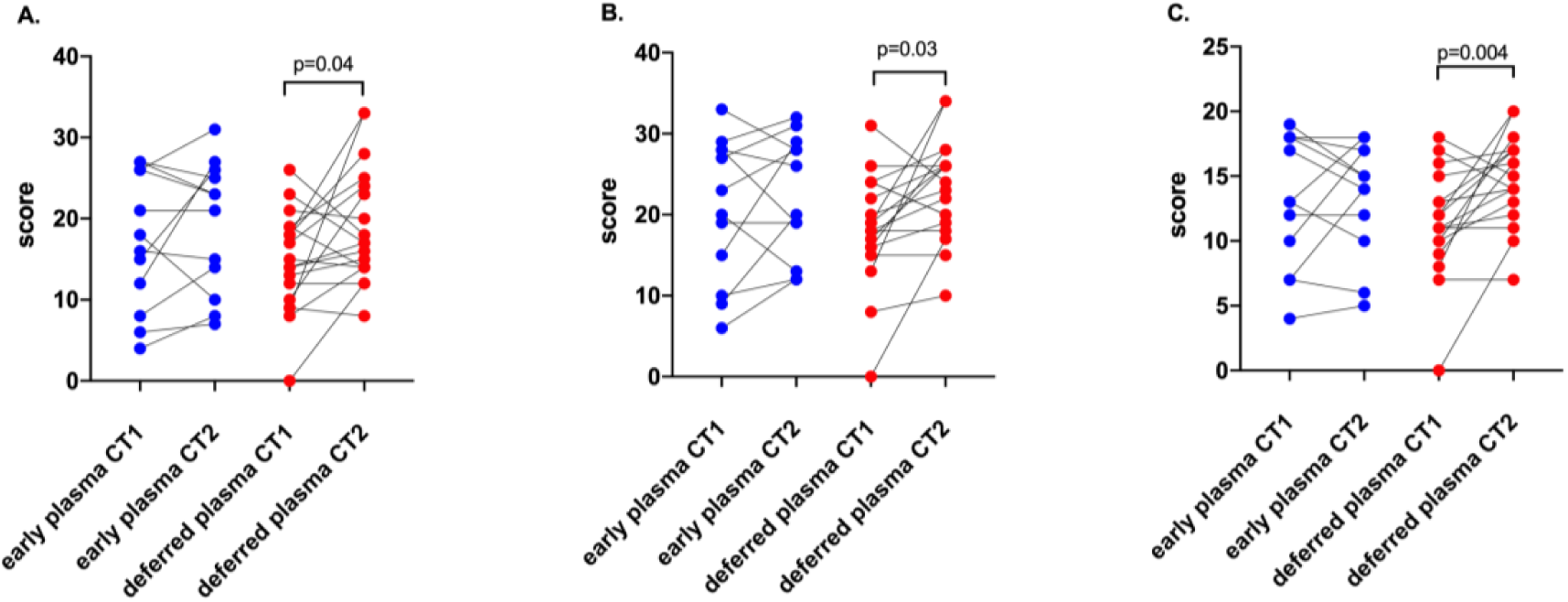
Chest CT COVID-19 pneumonia severity scores at baseline (day 0) and at day 5, for early convalescent plasma (n=12) and deferred plasma (n=18) group. A. CT score 1 (Zhou S. et al[20].) B. CT score 2 (Yang R. et al[23].) C. CT score 3 (Pan F. et al[21,22].)

### Immune response subgroup analysis

SARS-CoV-2 IgG titers were determined in patients who received early plasma and in the subset of patients from the deferred plasma group who had not yet received plasma, at baseline, day 3 and day 7. No significant differences were observed in IgG SARS-CoV-2 seropositive rate at the three time-point, nor in IgG seroconversion rates between plasma receptors and no plasma receptors at day 3 (69% vs 40%, p=0.07) or at day 7 (87 vs 83%, p=1.00) (Figure 5, A and B).

**Figure 5.**
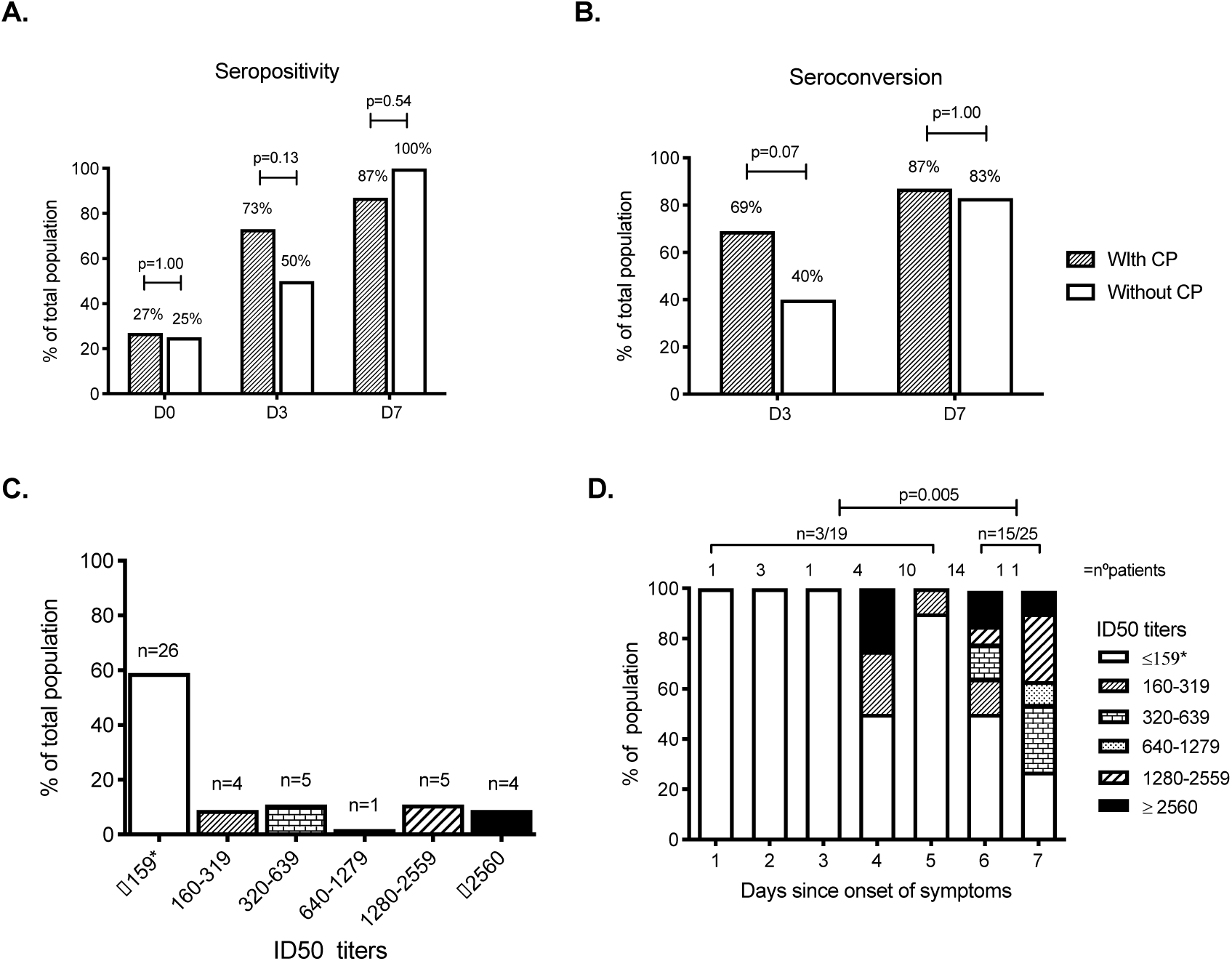
The humoral immune response induced by SARS-CoV-2. A. IgG seropositivity (OD ratio ≥ 1.1) analysis by sampling days. D0, day of enrollment; D3, the third day after enrollment; D7, seven days after enrollment. Dashes columns represent the patients who received convalescent plasma (CP) on enrollment (n=26 samples available on day 0, n=26 on day 3 and n=23 on day 7), white columns represent the patients from deferred plasma group that did not received plasma (n=20 samples available on day 0, n=20 on day 3 and n=12 on day 7). Above each column, the percentage of seropositivity is indicated. B. IgG seroconversion was considered if negative samples at 1:101 dilution increased to any positive dilution after 72h or 7 days, or if a 4-fold increase in end-point dilution titer from the enrollment was reached. Dashes columns represent patients who received CP, white columns represent patients from deferred plasma group that did not received plasma. Above each column, the percentage of seroconversion is indicated. C. Neutralizing antibody (NAb) titer measured by ID50 quantified at D0. The total number of patients reaching every dilution titer interval is indicated in each column. *ID50 titer ≤1:159 or no neutralization observed. D. NAb showed by day of COVID-19 symptoms onset. Each column represents the day after onset symptoms, above each column are the number of individuals, white painted represent individual with no NAb, painted columns represent different titers of NAb as shown in the figure. Capped lines represent Nab titers ≥1:160 before 5 days or after 6 days since the onset of symptoms.

Regarding NAbs titers, all infused plasma samples tested (n=18) had a positive ID50 at screening dilution titer (data not shown). The NAbs were also quantified for a total of 44 study patients: at enrolment, 59% (26/44) did not reach the screening cutoff (50% of neutralization) at the 1/160 dilution (Figure 5C). Interestingly, only 16% (3/19) of patients enrolled before day 5 since COVID-19 symptoms onset had ID50 titers ≥1:160, compared to 60% (15/25) of those enrolled after day 6 (p=0.005) (Figure 5D).

### Safety

Among all 41 patients receiving plasma in this study, there were four possibly related adverse events (3 cases of fever, 1 rash) and three SAEs (7.3%). Two patients developed severe respiratory deterioration within <6 hours after plasma infusion and were categorized as possible transfusion-associated acute lung injury (TRALI) type II[27]. One of the later patients additionally developed severe thrombocytopenia within 48h after plasma transfusion, with negative antiplatelet antibodies and megakaryocytic hyperplasia in the bone-marrow analysis. Platelet count remained low in the following weeks, despite platelet transfusions, steroids and immunoglobulin therapy, requiring splenectomy, rituximab and eltrombopag before slow stabilization. This event was diagnosed as a COVID-19 possibly related complication.

## Discussion

In this randomized clinical trial of symptomatic COVID-19 patients admitted early, we did not find significant differences in the composite primary outcome of death, mechanical ventilation or prolonged hospitalization, by administering immediate convalescent plasma, compared to plasma only in case of clinical worsening.

Convalescent plasma was not associated with a higher rate of SARS-CoV-2 RT-PCR clearance in nasopharyngeal swabs, suggesting that it does not provide a strong antiviral activity at this stage in patients with COVID-19. In accordance with this finding, transfused patients did not present a significant rise of SARS-CoV-2 IgG levels on days 3 and 7 compared to the natural increase in IgG titers in non-plasma infused patients, which could explain such lack of effect.

We actively selected patients at high risk of developing complications - based on CALL prediction score - and, over 40% of our participants developed severe respiratory failure. The lack of clinical benefit from convalescent plasma therapy in these patients may be explained by several reasons. Firstly, humoral immunity may not play a major role in the subset of patients who have already initiated a highly pro-inflammatory response and in whom inflammation and coagulopathy may be more important than viral replication[28]. We do not know whether pre-selection of plasma units with a very high concentration of NAbs could have succeeded in blunting this dysregulated inflammatory response. Additionally, an early adaptive immune response might be necessary to drive a more effective infection control. Indeed, different cellular and humoral responses are generated in mild or severe COVID-19 cases, and it has been reported that a specific cellular response can be detected early in the course of non-severe COVID-19[29],[30].

Secondly, limited efficacy may be due to a late administration in the course of the disease, in which a dysregulated immune response predominates and is independent of the virus cell entry blockade achieved by immunoglobulins[5,16]. Previous randomized trials of convalescent plasma for COVID-19 included patients who had a longer time gap between symptoms onset and transfusion as well as a more severe disease at enrollment[14,15]. Despite setting a strict ≤7 days of symptoms inclusion criterion, in our study, over 96% of subjects had already established pneumonia on enrollment. Hence, it is possible that some subjects had a more rapid or aggressive course or, particularly for older adults, true COVID-19 symptoms onset went unnoticed until several days into the disease course. Nonetheless, early-stage was well supported by the fact that on enrollment over 74% of our participants did not have detectable SARS-CoV-2 IgG, and about 60% did not have significant NAbs capacity.

Thirdly, given the design of our study, plasma administration in the deferred plasma group may have prevented the primary outcome from developing. However, the probability and time to progression into respiratory failure did not differ between both study groups. Since respiratory failure was the pre-specified criteria for plasma administration in the deferred plasma group, this secondary outcome allowed to compare early plasma versus no plasma, further supporting the lack of efficacy.

Plasma transfusion is not exempted from adverse events such as allergic reactions, infections transmission, and -very rarely-volume overload or TRALI[31]. In spite of the fact that the majority of clinical trials are still ongoing, convalescent plasma emergency use in COVID-19 treatment was recently approved by the FDA[32]. Reassuringly, in a recent report of 20,000 hospitalized patients receiving convalescent plasma for COVID-19, the incidence of related SAEs in the first four hours after infusion was <0.5%[33]. Nonetheless, in the present study, two participants developed acute respiratory failure after transfusion. Given that the patients were, according to the known evolution of COVID-19, in the peak of their inflammatory phase, it was challenging to determine if the respiratory failure corresponded to a TRALI[34].

Our study presents some limitations. Firstly, NAbs were not determined in donor plasma before the patient’s transfusion and we could not select plasma units with the highest neutralizing activity. Secondly, although we selected a very homogeneous population, it is possible that the study was underpowered to detect a statistically significant difference. Thirdly, as an open-label study, cointerventions such as steroid use may have unintendedly influenced outcomes[35]. Such management was not standardized, although alternative drug therapies were equally distributed in both study arms.

In conclusion, convalescent plasma transfusion in patients hospitalized in the early stage of COVID-19, compared to giving plasma only at clinical deterioration, did not improve clinical outcomes. Further research is needed to find the optimal use and timing of convalescent plasma in COVID-19.

## Data Availability

The full data that support the findings of this study are available from the corresponding author, upon reasonable request.

## Acknowledgments

We would like to thank all plasma donors who volunteered in great numbers and all patients who participated in the study. The institutional review board at Pontificia Universidad Católica de Chile for their efficient review of the application. All study nurses and in particular Haylim Nazar, Monique Moreau, Soledad Navarrete, Silvana Llevaneras, Carolina Henriquez and Camila Carvajal for their generous commitment to this project. The DIDEMUC for their collaboration in study finances management and coordination. Our colleagues Teresita Quiroga, and Ana María Guzmán for their collaboration with serologies and general laboratory analysis. All staff from the Diagnostic Virology Laboratory, Red de Salud UC CHRISTUS.

